# Molecular Epidemiology and evolutionary characteristics of dengue virus serotype-2 strains in Sri Lanka

**DOI:** 10.1101/2025.05.20.25328031

**Authors:** Dinuka Ariyaratne, Tibutius Thanesh, Pramanayagam Jayadas, Chandima Jeewandara, Bhagya Senadheera, Malithi De Silva, Laksiri Gomes, Farha Bary, Ayesa Syenina, Diyanath Ranasinghe, Heshan Kuruppu, Ananda Wijewickrama, Eng Eong Ooi, Gathsaurie Neelika Malavige

**Author notes:** Correspondence should be addressed to: Prof. Neelika Malavige DPhil (Oxon), FRCP (Lond), FRCPath, AICBU, Department of Immunology and Molecular Medicine, Faculty of Medical Sciences, University of Sri Jayewardenepura, Sri Lanka.

## Abstract

**Background:** Surveillance and characterization of different dengue virus (DENV) lineages of different serotypes causing outbreaks is crucial for initiation of timely dengue control measures and for implementing dengue vaccines. Therefore, we characterized the DENV-2 strains in Sri Lanka from 2016, until end of 2023 and their evolutionary dynamics to understand the geographical spread and mutations arising with the DENV-2 serotypes in Sri Lanka

**Methodology:** Sequencing was carried out on 80 DENV-2 samples collected from patients with acute dengue recruited from 2016 to 2018 and on 12 DENV-2 samples from 2022 to 2023 using Oxford Nanopore Technology. Phylogenetic analysis was carried out using the IQ-TREE tool within Galaxy to construct a phylogenetic tree from the aligned sequence data. The sequences were also analyzed for non-synonymous amino acid changes in the envelope and NS1 regions.

**Results:** The Sri Lankan DENV-2 sequences circulating from 2016 to end of 2023 were found to belong to genotype II.F.1 lineage. They were closely related to strains circulating during the same period in South Asia and Southeast Asia. We identified 15 non-synonymous mutations within the envelope region and 22 non-synonymous mutations within the NS1 region, with 7 non-synonymous mutations within the E region (M6I, Q52H, E71A, V129I, N390S, I484V, T478S) and 10 non-synonymous mutations within the NS1 region (S80T, T117A, Q131H, K174R, F178S, N222S, L247F, I264T, T265A, K272R) seen in all sequenced samples. Some of these mutations were previously shown to be associated with increase in viral replication, NS1 secretion and immune evasion, while some have not been reported elsewhere.

**Conclusions:** Given the increase in dengue transmission in many countries, it is important to further strengthen DENV surveillance for studying the evolutionary patterns of the DENV to initiate timely and appropriate control measures.

## Background

There has been an alarming rise of dengue infections in all regions of the world, with year 2024 reporting the highest number of dengue cases so far [1]. 3.83 billion individuals (over 53% of the global population), is estimated to be living in areas that are suitable for dengue transmission and this is expected to rise further in future due to climate change [2]. Over 13 million cases were reported globally by August 2024, though the actual number is likely to be higher due to under reporting and the difficultly in clinically differentiating dengue from many other similar febrile infections[1, 3]. Yet, the number of dengue infections is forecasted to keep rising every year while spreading to new geographical regions due to multiple factors such as climate change, rapid urbanization, population displacements and possibly rapid evolution of the dengue virus (DENV)[4]. As a result, the World Health Organization has included it in the R&D Blueprint as an emerging pathogen of pandemic potential [5].

Dengue infections are caused by four related but genetically distinct dengue virus serotypes (DENV1 to 4), which share 60 to 70% homology with each other [6]. Each DENV serotype consists of multiple lineages with a nucleotide diversity of 6 to 8% [7]. Although DENVs are endemic in many countries in the tropical and subtropical regions, major shifts in the circulating DENV serotype leads to large outbreaks, overwhelming health care systems [8–11]. Moreover, due to increased transmission rates, many countries have reported co-circulation of multiple DENV serotypes and lineages resulting in outbreaks [12–14]. Different DENV lineages have been shown to result in different magnitudes of infectivity, viral titers and pathogenicity in *in vitro* models [15], with certain lineages known to cause more severe infection and larger outbreaks [16, 17]. These phenotypic differences between the DENV lineages may be attributed to sequence-dependent virus-host interactions. For instance, a single nucleotide substitution in an American DENV2 genotype may explain the lack of outbreak in Tonga in contrast to large number of severe dengue cases elsewhere in the Pacific during the early 1970s [18] Moreover, immunity from dengue vaccination and the use of Wolbachia mosquitoes in vector control may impose new selection pressures on DENV, thus creating the possibility of new virus lineages. Therefore, surveillance and characterization of circulating DENV lineages of different serotypes would be important to understand how they may affect clinical disease severity and transmission dynamics. In this study, we characterized the DENV-2 strains in Sri Lanka from 2016, until end of 2023 and their evolutionary dynamics to understand the geographical spread and mutations arising with the DENV-2 serotypes in Sri Lanka.

## Methods

### Recruitment of patients with acute dengue infection for serotyping PCR and DENV sequencing

Blood samples were collected from 582 adult patients who were clinically suspected to have an acute dengue infection, following informed consent during the period of July 2016 to October 2018, and again 467 adult patients were recruited between November 2022 to December 2023, in the National Institute of Infectious Diseases (NIID), Angoda, Sri Lanka. Quantitative real time PCR for detection of the four DENVs were carried out in all samples to determine the infecting DENV serotype using modified CDC primers as previously described[19].

### Ethical Approval

Ethics approval for the study was obtained from the Ethics Review Committee, University of Sri Jayewardenepura and the administrative clearance was obtained from the Ministry of Health, Sri Lanka. All individuals gave informed written consent.

### Whole genomic sequencing of the DENV2 that circulated in Sri Lanka during 2017 to 2018 and 2023

As DENV-2 was the predominant circulating serotype since mid-2016 and was responsible for over 95% of infections in 2017[16], 80/582 DENV2 positive samples from mid-2016 to end of 2018 and 12/467 DENV2 positive samples from November 2022 to December 2023 were selected for sequencing using an amplicon-based framework [20] All samples selected for sequencing passed the cycle threshold value of <25. Briefly, cDNA was synthesized from viral RNA using the SuperScript IV First-Strand Synthesis System (Life Technologies) – for the 80 samples collected from 2016-2018, or Lunascript (LunaScriptTM RT SuperMix Kit, New England Biolabs, E3010) – for the 12 samples collected between 2022-2023. Amplicons were then generated using Q5 Hot Start High-Fidelity DNA Polymerase (New England Biolabs) with DENV-2 specific primers. The PCR mixture was initially incubated for 30s at 98 ◦C for denaturation, followed by 35 cycles of 98 ◦C for 15 s and 65 ◦C for 5 min. The PCR products were then diluted 10-fold. Subsequently, the diluted amplicons were end-prepped and dA-tailed using NEBNext Ultra II End Repair/dA-Tailing Module. Native barcodes and sequencing adapters supplied in the EXP-NBD101/114 kit (Oxford Nanopore Technologies) were attached to the dA-tailed amplicons using NEB Blunt/TA Ligase 2x Master Mix and NEBNext Quick Ligation Kit, respectively. Finally, 15 ng of DNA library was loaded on to the R9.4.1 flow cell following the SQK-LSK109 ligation sequencing kit (Oxford Nanopore Technologies) protocol, the sequencing run was performed on the Minion Mk1b (Oxford Nanopore Technologies) for a total of 24 hours. Reads were basecalled using the MinKnow software.

### Bioinformatics analysis

The sequences were analyzed and compared with other DENV-2 sequences from Sri Lanka collected in 1996 (GenBank FJ882602.1) and 2003 (GenBank GQ252677.1), accessed from GISAID. The initial alignment of the FASTA files was done with MAFFT and the phylogenetics tree was built using IQTree. The sequences were then analyzed for non-synonymous amino acid changes in the envelope and NS1 regions. Out of these non-synonymous changes, we selected the amino acid mutations unique to the 2016/2017/2018 and 2022/2023 sequences compared to the reference genome (GenBank NC001474.2)

### Generation of Maximum-Likelihood Tree

After generating FASTA files for all sequences sequenced in 2017 to 2023, sequences with more than 70% coverage overall and 100% coverage of E and NS1 regions were selected (n=63). They were aligned with the reference sequence NC 001474.2 and previous Sri Lankan DENV-2 sequences from 1996 (GenBank FJ882602) and 2003 (GenBank GQ252676) using MAFFT [21]by running on the public server of the web platform Galaxy (http://usegalaxy.org).

These aligned sequences were run in Nextclade [22] website (https://clades.nextstrain.org) to classify the genomes based on the latest classification [7](https://dengue-lineages.org/)and to analyze the non-synonymous mutations and resulting amino acid changes in the E andNS1 regions. Publicly available DENV-2 sequences (n=362) were downloaded from NCBI Virus [23]database. The sequences were filtered to obtain complete whole genome sequences from Asia and South America between 01.01.1994-28.10.2024. These sequences (n=425) were aligned with the previous sequences and redundant sequences were removed. The aligned sequences were then used to build a maximum-likelihood tree (1000 bootstraps) using IQ-Tree[24]. The tree was then visualized in the online tree viewing platform Interactive Tree of Life [25] (https://itol.embl.de/), colour coded and annotated.

### Molecular Clock Analysis

We undertook a comprehensive approach for molecular clock analysis of DENV-2, starting with sequence collection and preparation. Initially, we gathered a total of 63 DENV-2 sequences from Sri Lanka, which were organized in FASTA format. To supplement this dataset, we retrieved additional DENV-2 sequences from the National Center for Biotechnology Information (NCBI) database, specifically focusing on sequences from human hosts with a minimum length of 10,000 nucleotides. We excluded sequences from environmental sources and laboratory-passaged sequences, resulting in the collection of 754 sequences.

Next, we standardized the FASTA headers across both the Sri Lankan and NCBI datasets using a Python script to ensure consistency, adopting the format: >Reference_Description_Date. These sequences were then uploaded to the Galaxy platform using its “Upload Data” tool for data processing, where we merged them into a single FASTA file with the “Concatenate FASTA” tool, culminating in a combined dataset of 817 sequences. Subsequently, we performed multiple sequence alignment using MAFFT in Galaxy, and any sequence lacking a recorded collection year was eliminated from the dataset. To enhance clade annotation, we uploaded the aligned sequences to Nextclade, which provided clade information that was then downloaded in Excel format. We created a comprehensive metadata CSV file containing sequence names, collection dates, and clade information, ensuring that any sequence missing date or clade information was excluded.

For phylogenetic analysis, we employed the IQ-TREE tool within Galaxy to construct a phylogenetic tree from the aligned sequence data, subsequently downloading the tree in Newick format. The resulting phylogenetic tree file, along with the metadata CSV, was utilized as input for Clockor2[26] to conduct the molecular clock analysis. Clockor2 facilitated root-to-tip regression analysis, allowing us to infer both global and local strict molecular clocks. During this analysis, it was observed that NCBI sequences belonging to clade 2I exhibited a downward regression trend, prompting their removal to ensure accurate molecular clock estimation. The decision to exclude these sequences was based on their deviation from expected evolutionary rates, which could potentially introduce bias into the molecular clock inference.

### Geospatial Mapping of Dengue Serotype 2 lineages

For visualizing the geographical distribution of DENV-2 lineages, we leveraged a geospatial visualization approach utilizing Python’s geospatial data libraries. First, we loaded the metadata of DEN-2, which included information on sequence lineages and their locations, from a TSV file into a Pandas DataFrame. This metadata file was downloaded from the Nextstrain platform for comprehensive genomic analysis of DENV-2.

To map these data points, we utilized a Natural Earth shapefile representing world countries at a high resolution (10m scale). This shapefile was loaded as a GeoDataFrame using the Geopandas library, ensuring that the geographical information of each country could be effectively utilized in plotting. Certain location names in the metadata, like ‘California (USA)’ and ‘Florida (USA)’, were standardized to ‘United States of America’ to ensure accurate mapping. We then merged the metadata DataFrame with the GeoDataFrame of world countries based on location names, aligning DENV-2 lineage data with their respective geographical geometries.

The resulting GeoDataFrame was filtered to retain only valid geometries, and centroid calculation was performed for plotting purposes. We utilized Matplotlib to initialize a plot, setting the world map boundaries with a light grey color. The DENV-2 lineage points were overlaid on this map, color-coded according to lineage, using the HSV colormap for differentiation.

### Statistical Analysis

R (version 4.4.1) using R packages stats, dplyr, ggplot2, ggpubr, tidyverse,ggsignify, patchwork, pheatmap. K-means clustering was conducted using R with the stats package. We chose k = [3] based on the elbow method. The kmeans function was then applied with nstart = 20 to minimize the risk of local optima. All the R and Python code used for analysis and figure generation can be accessed at: https://github.com/AICBU/DENV2_SriLanka.

## Results

### Full-length genomic sequence of the circulating DENV-2 in Sri Lanka

The largest dengue outbreak in Sri Lanka was happened in 2017 due to the emergence of DENV-2 resulting in 186,101 hospital admissions [16]. Whole genome sequencing of DENV2 positive samples collected during the outbreak between 2017-2018 found the circulating DENV2 strain to be genotype II, major lineage F and minor lineage 1 (genotype II.F.I) based on the latest classification of DENVs (Figure 1)[7]. Phylogenetically, the 2017/18 sequences were closely related to strains circulating during the same period in China and other Asian countries such as Thailand, Singapore and Malaysia (Figure 1). DENV2 positive samples were also collected between 2022-2023. The whole genome sequencing on these samples revealed they were of the same genotype II F1.1 strain as the 2017/2018 samples. The 2017/2018 and 2022/2023 sequences formed a new DENV-2 genotype II.F.I clade, clustering with other previously reported 2017 Sri Lankan DENV-2 strains (MT180479, GenBank)[27], and genetically distant from the previous DENV-2 strains that circulated in Sri Lanka from 1996 to 2003, which belonged to DENV-2 genotype II_A (Sup Figure 1). These DENV-2 strains from 1990s and early 2000s belong to the major lineage A, which was mainly reported from Africa in the 1990s, which has since evolved to II_A1 by 2007 and subsequently II_A2.2 by 2018 (Figure 2). The more recent F1.1 lineage is predominantly spread in East and Southeast Asia (Figure 2).

**Figure 1:**
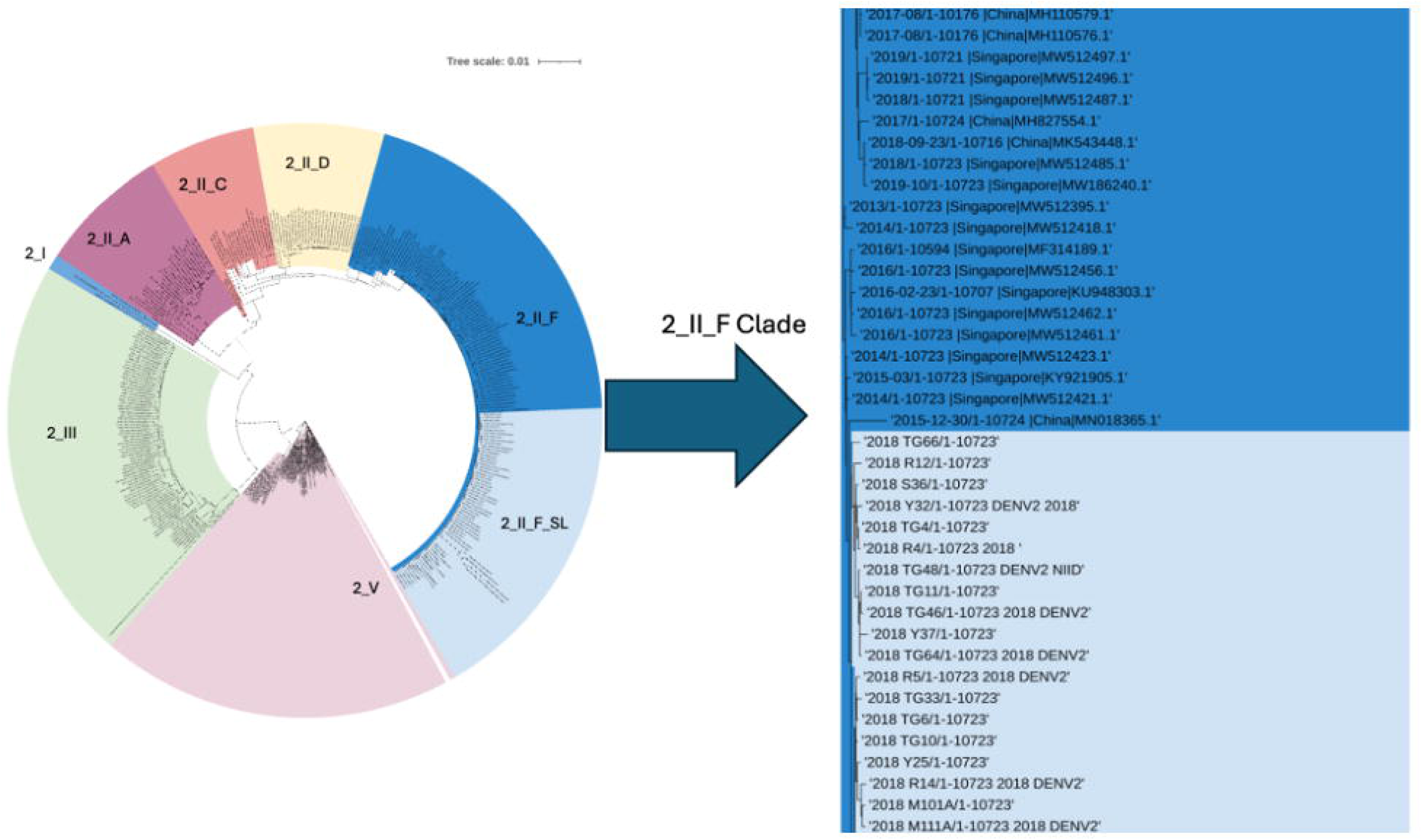
The phylogenetic analysis of DENV2 sequences in Sri Lanka. The phylogenetic tree was generated with the Sri Lankan DENV2 strains in comparison to a randomized sub-selection of DENV2 serotype 2 strains reported in Asia and South America in the last 30 years (1994-2024). The Sri Lankan DENV2 strains from 2016 onwards (highlighted light blue) cluster together with genotype II, major lineage F strains (dark blue) from East and Southeast Asia, reported in 2013-2018. Genotype II, major lineage A is indicated in red, major lineage C indicated in orange, major lineage D in yellow, genotype 1 in cyan, genotype III in green and genotype V in purple.

**Figure 2:**
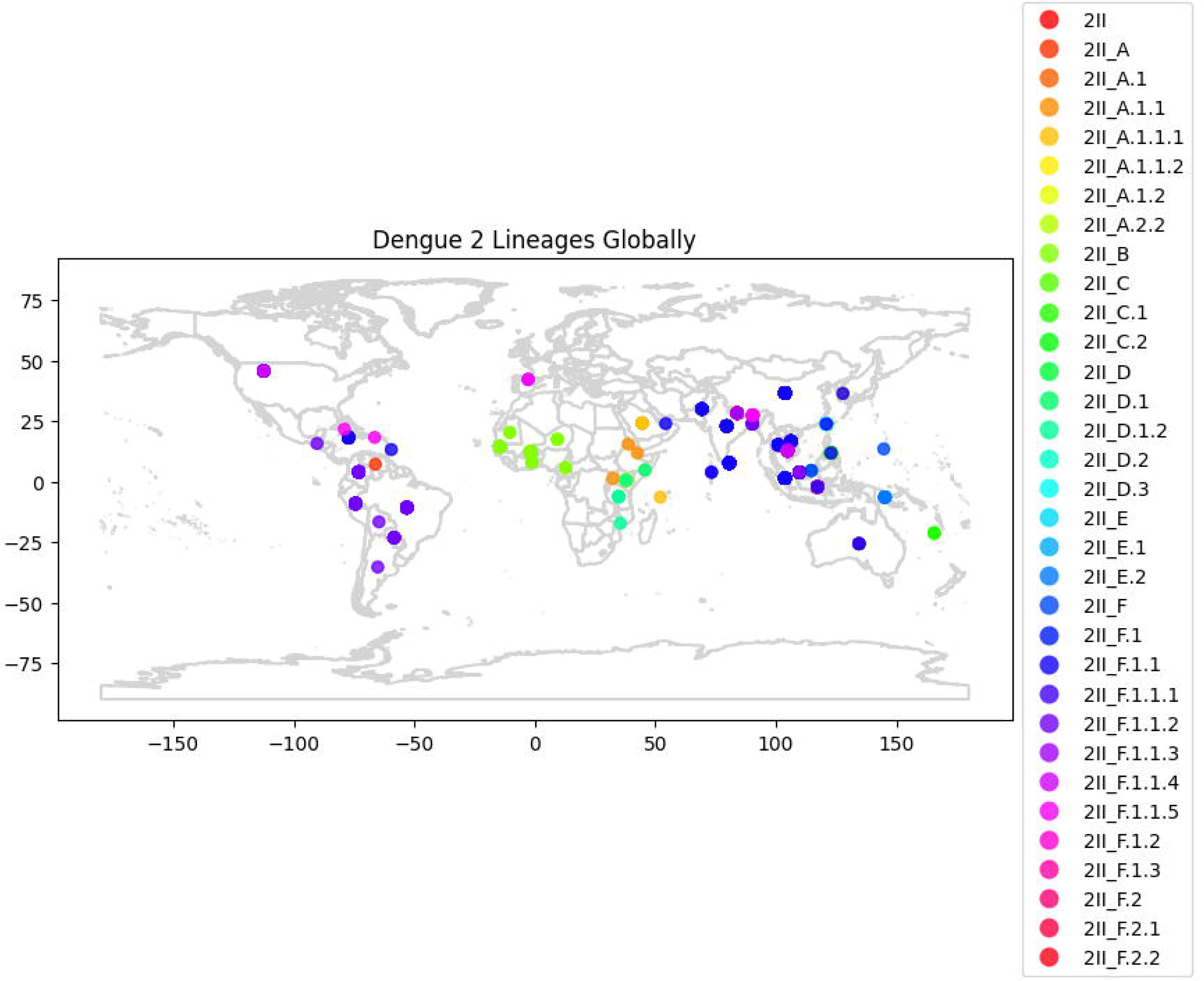
Geographical distribution of Dengue 2 Genotype 2 Lineages. An illustration of the spatial distribution of the different DENV-2 genotype 2 lineages across the world. Centroids, colour coded according to the major and minor lineages (right-hand-side panel) appear in the geographical locations they have been reported from. Data was accessed from Nextstrain[22].

### Evolution of DENV-2 genotype II. F.I strains in Sri Lanka

The Sri Lankan DENV-2 strains that were responsible for the large outbreak in 2017, gradually evolved from 2016 to 2023, with the sequences falling along regression line as shown in the molecular clock analysis (Figure 3). The Sri Lankan DENV-2 genotype II. F.I strains thus appeared to evolve at a similar rate (1.445 × 10^-3^) compared to evolutionary rates of other reported DENV2 strains globally (1.45× 10−3) (Supplementary Table 1)[28].

**Figure 3:**
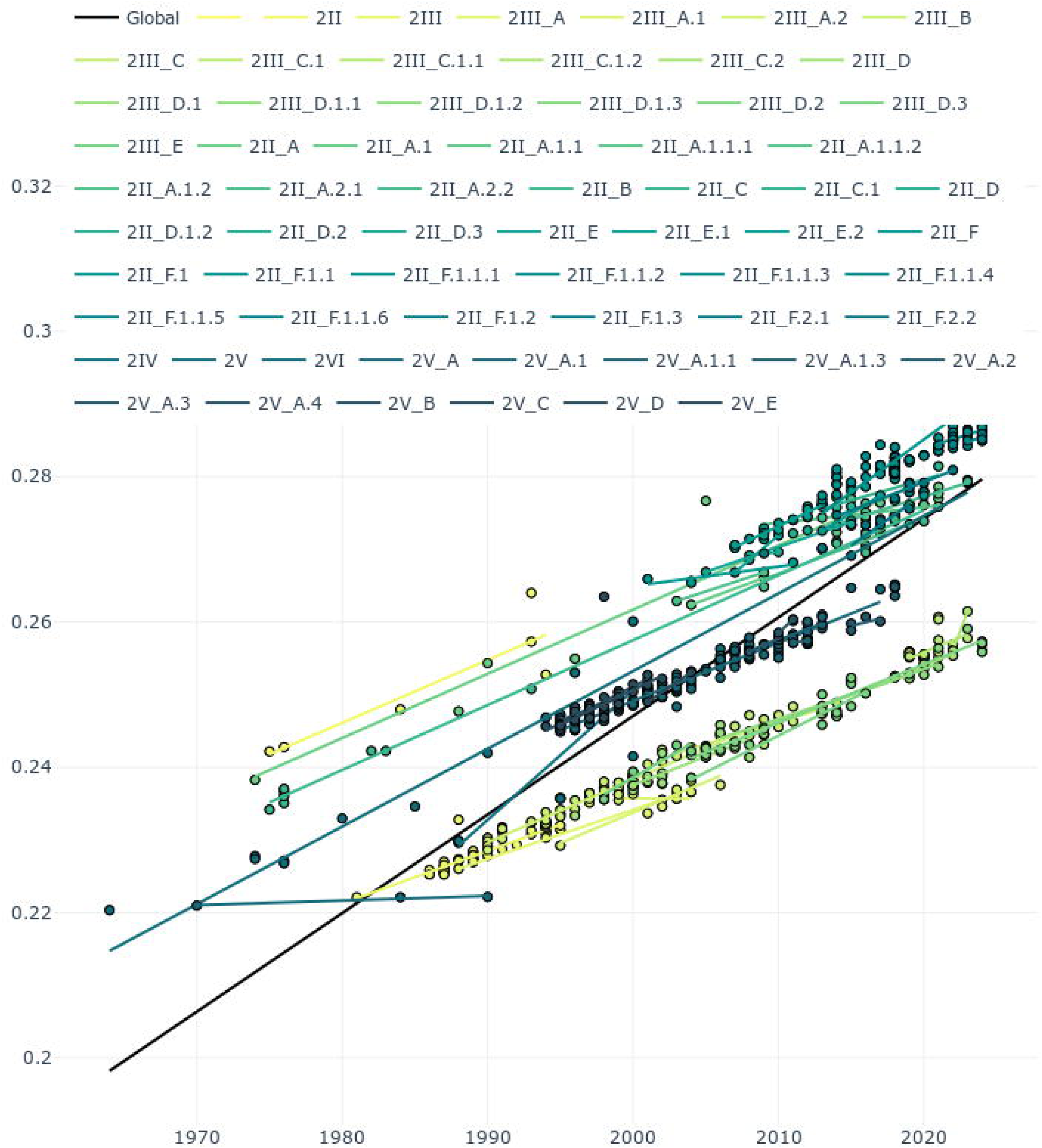
Phylogenetic Tree of Dengue Virus Serotype 2 (DENV-2) Derived from Molecular Clock Analysis. A time-scaled phylogenetic tree was constructed (n=768 sequences) using the Maximum Likelihood method, illustrating the genetic relationships and divergence timing among DENV-2 strains collected from Sri Lanka (n=63) and various global locations (n=705). Each branch is scaled according to the molecular clock, with branch lengths proportional to the estimated amount of evolutionary time elapsed. Clockor2 [26] was used to perform root-to-tip regression analysis to infer global and local strict molecular clocks Significant nodes are highlighted with asterisks, denoting posterior probabilities greater than 0.95, indicating strong support for these clades. Scale bar indicates nucleotide substitutions per site. Colors (green shades) represent different geographic origins as mentioned in the key above the tree. Global Line (Black Line). In this molecular clock, a generally linear trend is observed, supporting a clock-like evolution.

### Characterization of mutations specific to E and NS1 in DENV-2 genotype II.F.I

Analysis of the mutations within DENV-2 genotype II.F.I compared to the reference genome (GenBank: NC 001474.2) showed 15 non-synonymous mutations within the envelope (E) region and 22 non-synonymous mutations within the NS1 region, occurring with varying frequencies (Supplementary Table 2). There were 7 non-synonymous mutations within the E region (M6I, Q52H, E71A, V129I, N390S, I484V, T478S) and 10 non-synonymous mutations within the NS1 region (S80T, T117A, Q131H, K174R, F178S, N222S, L247F, I264T, T265A, K272R) seen in all 48 samples. These non-synonymous mutations within the E protein were seen in the fusion loop (one), EDI (two), EDII (one), EDIII (one) and transmembrane region (two). All sequenced DENV-2 genotype II.F.I strains shared a “core” set of ELprotein and NS1Lprotein changes (Figure 4). Some of the newer substitutions, such as the W453C in E proteins or the F279L mutation in the NS1 protein are not seen in the DENV-2 genotype II.F.I strains in 2017/2018 but only appear in the strains sequenced during the 2022 to 2023 period (Figure 4). However, due to the relatively small sample size of a total of 63 sequences used in this study, how these different sub-lineages arising out of the DENV-2 genotype II.F.I associated with clinical disease outcomes could not be analysed.

**Figure 4.**
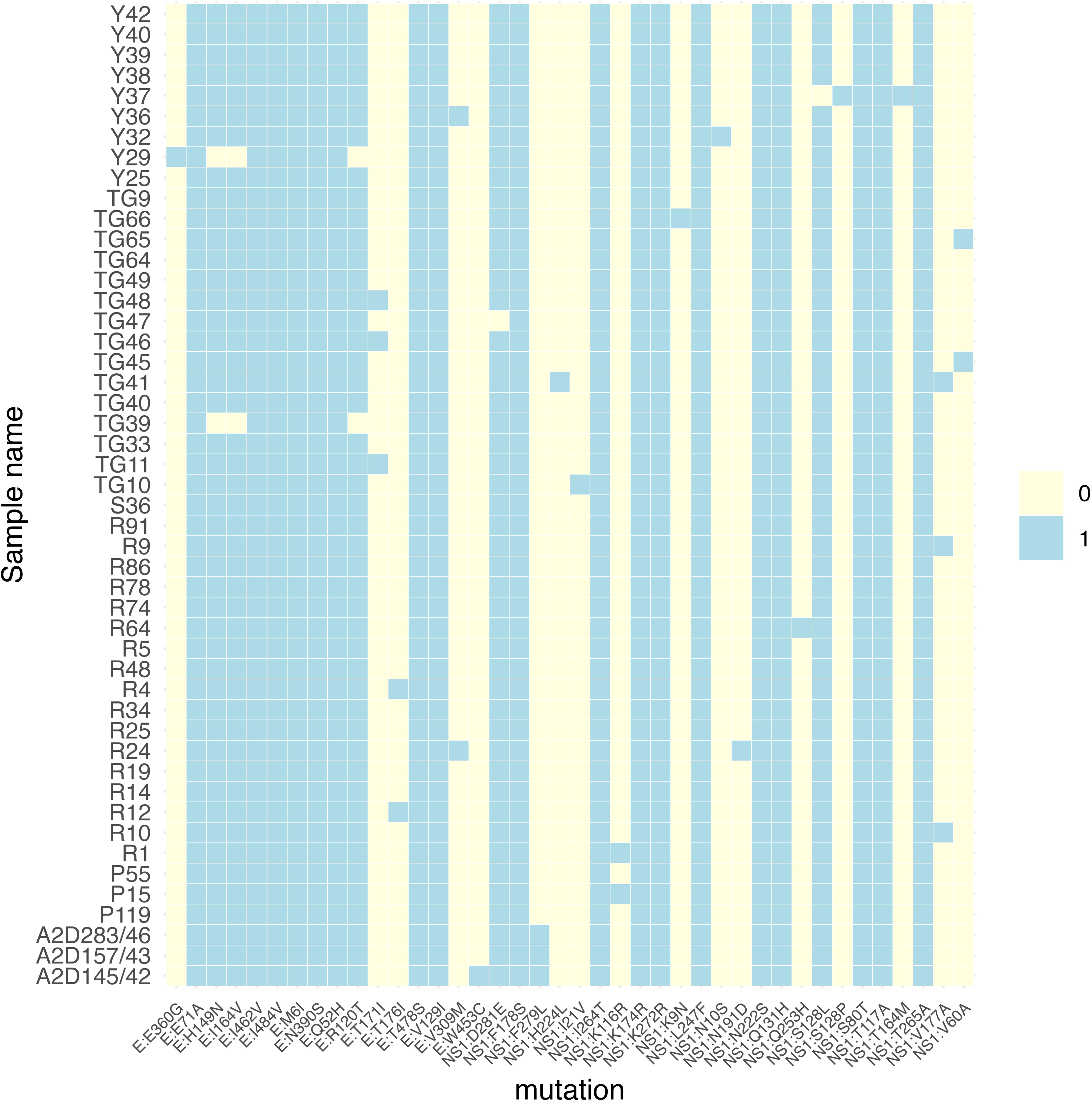
A heatmap of non-synonymous mutations within the envelope and NS1 regions identified in the DENV-2 genotype II.F.I strains from Sri Lanka. Rows correspond to individual samples, which are in chronological order from top to bottom (2016 to 2023) and columns correspond to specific E: or NS1: amino acid substitutions. Mutations are in the order of their positions in the E and NS1 region. A light-yellow cell indicates the absence (0) of the indicated mutation, while a light-blue cell indicates its presence (1). The color bar to the right shows the binary scale for presence and absence. The 2022/2023 samples at the bottom show 3 new mutations not seen in the samples sequenced from 2016 to 2018.

## Discussion

Outbreaks due to DENVs have been reported in Sri Lanka since the 1960s, while dengue outbreaks resulting in dengue haemorrhagic fever (DHF) only occurred since 1989 [8]. Since all four DENV serotypes had been circulating in Sri Lanka with DENV3 and DENV2 serotypes co-circulating and causing outbreaks until year 2009 [8, 29]. In this study we show that the introduction of a new DENV-2 genotype in mid-2016, were responsible for the largest dengue outbreak in Sri Lanka in 2017. Interestingly, although the outbreaks with this DENV2 genotype reduced in subsequent years, it was found to become endemic and continue to cause outbreaks in Sri Lanka, even during the COVID-19 pandemic [30]. Although these DENV-2, genotype II strains were initially classified as belonging to the cosmopolitan genotype [16], based on the earlier DENV genotype classification, using the new proposed classification, they were found to form a new lineage of genotype II.F.I [7]. Therefore, although the cosmopolitan genotype of DENV-2 was reported from all regions in the world [17], DENV-2 genotype II.F.1 was only reported in South Asia and Southeast Asia. This shows how the new DENV classification provides more granularity regarding the evolution and geographical spread of the DENV genotypes as it enables characterization of sub-lineages of different genotypes.

The DENV-2 genotype II.F.I strains in Sri Lanka showed a similar evolutionary rate as other DENV-2 strains [28]. We observed 15 non-synonymous mutations within the envelope region and 22 non-synonymous mutations within the NS1 region in this strain. Three mutations within the envelope region (E71A, H149N and N390S) have been previously described as mutations unique to the DENV-2 cosmopolitan genotype [17]. In addition to these three previously described mutations, the four other new mutations seen in all Sri Lankan strains were seen in domains I, II, III and the transmembrane region of the protein. Therefore, it would be important to study further if these mutations could affect the binding of neutralizing antibodies and thereby possibly leading to re-infection with DENV-2 as described previously [31, 32]. Further, all sequenced DENV-2 strains from 2016 to end of 2023 showed 10 non-synonymous mutations within the NS1. Although these mutations have not been previously described as being unique to the cosmopolitan strain, some of these mutations have shown to be associated with pathogenicity. For instance, the T117A was shown to produce 12-fold more intracellular virus than the wild-type strains [33]. The K272R substitution in NS1, which was identified in the DENV-2 virus that led to a large outbreak in 2015 in Taiwan, was shown replicate faster, produce more secretory NS1 in cell culture supernatants and inhibited STAT1 phosphorylation thereby inhibiting interferon signaling pathways [34]. While some other mutations observed by us have been described in other DENV-2 genotypes [15], DENV-2 genotype III in Brazil [35] and in other genotypes [27], it would be important to further study the implications of these mutations in transmission of the virus and altering its pathogenicity.

The DENV-2 genotype II.F.I strains in Sri Lanka had four new mutations in the envelop region and 10 non-synonymous mutations within the NS1, which are not found in any of the circulating DENV-2 viruses elsewhere. Therefore, it is possible that these genetic changes within DENV-2 genotype II.F.I strains could at least partially contribute to the change from this genotype transitioning from epidemic to endemic transmission. Although factors that lead to evolution and adaptation of the DENV are not entirely clear, it is likely that these mutations could have been driven by either human or Aedes hosts in Sri Lanka. As dengue case numbers in Sri Lanka report patients hospitalized with clinically suspected dengue infection, the actual infections dengue infections are very much underestimated. Importantly, asymptomatic infection is also known to cause transmission [36], and it could be the main selection factor for development of mutations to adapt to acquiring endemicity. As dengue pathogenesis is underpinned largely by virus-host interactions, local population genetic factors both of humans and Aedes may drive geographic dependent nuances in DENV diversity. The impact of such DENV diversity on dengue across the different ethnic groups living in the dengue endemic world should be investigated promptly.

The genetic diversity between different genotypes of DENV serotypes have also shown to affect vaccine efficacy. For instance, the CYD-TDV phase III vaccine trial showed different DENV2-specific vaccine efficacy in Asian and Latin American populations [37, 38]. These differences may, at least in part, have been influenced by the antigenic distance between the vaccine strain and circulating DENV reduced CYD-TDV vaccine efficacy across study sites[39, 40]. Therefore, given that the strains used to develop the currently licensed dengue vaccines composed of historical DENV strains, it is important to continuously carry our surveillance of different DENV strains in different parts of the world.

## Conclusions

In summary, in this study we have characterized the DENV-2 strains from 2016 to end of 2023, which were responsible for the largest ever dengue outbreak in Sri Lanka, which were found to be genotype II.F.I. The Sri Lankan DENV-2 strains were found to have certain mutations which have been associated with increase in viral replication, NS1 secretion and immune evasion and several unique mutations, which may have contributed to transition from epidemic transmission to endemicity in Sri Lanka. Given the increase in dengue transmission in many countries, it is important to further strengthen DENV surveillance for studying the evolutionary patterns of the DENV to initiate timely and appropriate control measures.

## Data Availability

All data is available within the manuscript and the supporting files.

## Abbreviations

CDC: Centers for Disease Control
DENV: Dengue virus
RNA: Ribonucleic acid
STAT1: Signal transducer and activator of transcription 1

## Declarations

## Ethics approval and consent to participate

Ethics approval was obtained from the Ethics Review Committee of the University of Sri Jayewardenepura. All participants gave informed written consent.

## Consent for publication

Not applicable.

## Availability of data and material

All data is available within the manuscript and the supporting files.

## Authors’ contributions

Conceptualization: DA, GNM, TTPJ

Data curation: DA, HK, LG, TTPJ, AW

Experiments and investigations: DA, TTPJ, HK, LG, FB, AS

Data analysis: BS, MDS, DR

Project administration and supervision: GNM, CJ, EEO, AW

Funding acquisition: GNM, CJ, EEO

Writing the manuscript: DA, GNM, EEO

Reviewing the manuscript: EEO

---

**Supplementary Figure 1:** Phylogenetic analysis of DENV2 genotype II strains. The phylogenetic tree was generated with 152 DENV2 genotype II strains (global and Sri Lankan) with the major lineages A, B,C,D,E,F denoted in different colours indicated in the colour key on the left side. The right inset shows the F major lineage with the Sri Lankan sequences (white font) and the global sequences they cluster with.

