## Supplementary figures and images for "Molecular Epidemiology and evolutionary characteristics of dengue virus serotype-2 strains in Sri Lanka"

### Supplementary figures

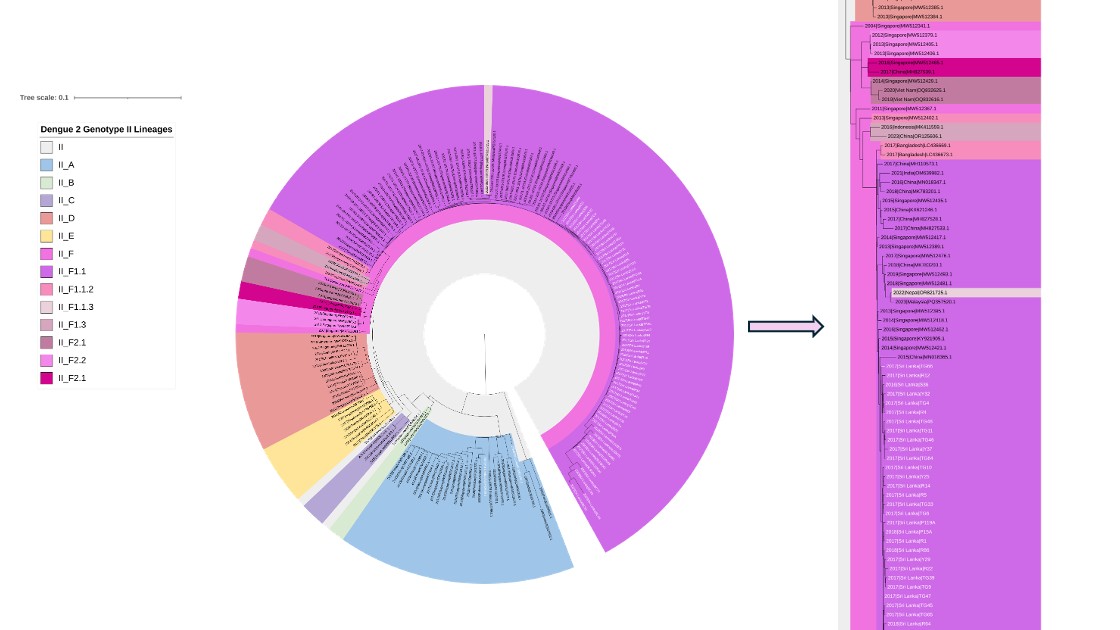
