## Supplementary data for "Molecular Epidemiology and evolutionary characteristics of dengue virus serotype-2 strains in Sri Lanka"

**Supplementary Tables**

**Supplementary Table 1**

| **Dengue Protein** | **Non-synonymous mutation** | **Region** | **Frequency** | **Percentage** |
| --- | --- | --- | --- | --- |
| Envelope | E:Q52H | Domain I | 48/48 | 100.00 |
| Envelope | E:M6I | Domain I | 48/48 | 100.00 |
| Envelope | E:E71A | Fusion loop | 48/48 | 100.00 |
| Envelope | E:R120T | Domain II | 46/48 | 95.83 |
| Envelope | E:V129I | Domain II | 48/48 | 100.00 |
| Envelope | E:H149N | Domain I | 46/48 | 95.83 |
| Envelope | E:I164V | Domain I | 46/48 | 95.83 |
| Envelope | E:T171I | Domain I | 3/48 | 6.25 |
| Envelope | E:T176I | Domain I | 2/48 | 4.17 |
| Envelope | E:V309M | Domain III | 2/48 | 4.17 |
| Envelope | E:E360G | Domain III | 1/48 | 2.08 |
| Envelope | E:N390S | Domain III | 48/48 | 100.00 |
| Envelope | E:W453C | Transmembrane | 1/48 | 2.08 |
| Envelope | E:I484V | Tansmembrane | 48/48 | 100.00 |
| Envelope | E:T478S | Transmembrane | 48/48 | 100.00 |
| NS1 | NS1:K9N | Beta roll | 1/48 | 2.08 |
| NS1 | NS1:N10S | Beta roll | 1/48 | 2.08 |
| NS1 | NS1:I21V | Beta roll | 1/48 | 2.08 |
| NS1 | NS1:V60A | Wing | 2/48 | 4.17 |
| NS1 | NS1:S80T | Wing | 48/48 | 100.00 |
| NS1 | NS1:K116R | Wing | 2/48 | 4.17 |
| NS1 | NS1:T117A | Wing | 48/48 | 100.00 |
| NS1 | NS1:S128L | Wing | 47/48 | 97.92 |
| NS1 | NS1:S128P | Wing | 1/48 | 2.08 |
| NS1 | NS1:Q131H | Wing | 48/48 | 100.00 |
| NS1 | NS1:T164M | Wing-connector | 1/48 | 2.08 |
| NS1 | NS1:K174R | Wing-connector | 48/48 | 100.00 |
| NS1 | NS1:V177A | Wing-connector | 3/48 | 6.25 |
| NS1 | NS1:F178S | Wing-connector | 48/48 | 100.00 |
| NS1 | NS1:N191D | Beta ladder | 1/48 | 2.08 |
| NS1 | NS1:N222S | Spaghetti loop | 48/48 | 100.00 |
| NS1 | NS1:H224L | Spaghetti loop | 1/48 | 2.08 |
| NS1 | NS1:L247F | Spaghetti loop | 48/48 | 100.00 |
| NS1 | NS1:Q253H | Spaghetti loop | 1/48 | 2.08 |
| NS1 | NS1:I264T | Spaghetti loop | 48/48 | 100.00 |
| NS1 | NS1:T265A | Spaghetti loop | 48/48 | 100.00 |
| NS1 | NS1:K272R | Spaghetti loop | 48/48 | 100.00 |
| NS1 | NS1:F279L | Spaghetti loop | 3/48 | 6.25 |
| NS1 | NS1:D281E | C terminus | 47/48 | 97.92 |

**Supplementary Table 01: Non-synonymous mutations of DENV2 sequences in Sri Lanka.** The non-synonymous mutations in the envelope(E) and non-structural protein 1 (NS1) and their frequencies of occurrence in the 48 samples sequenced. The regions within the envelope and NS1 are marked according to the structures previously described [1, 2]

**Supplementary Table 02**

| **Clade** | **Tips** | **Rate** | **R-squared** |
| --- | --- | --- | --- |
| 2II_A | 7 | 8.841 x 10^-4 | 0.907 |
| 2II_A.1.2 | 2 | 6.532 x 10^-4 | 1 |
| 2II_F.1.1 | 99 | 1.445 x 10^-3 | 0.438 |
| 2V | 19 | 1.040 x 10^-3 | 0.963 |
| **Global** | **768** | **1.328 x 10^-3** | **0.7** |

**Supplementary Table 2:** Evolutionary rates of DENV2 global and Sri Lankan sequences based on molecular clock analysis. The evolutionary rates of global DENV2 and the rates of different genotypes and lineages (2_IIA, 2_IIF, 2_IIV) seen in Sri Lanka between 1996-2024 were analyzed using a molecular clock method. The R2 value being closer to 1 is indicative of a good fit to the molecular clock model for this clade, indicating that the rate is consistent over the specified timeline.
